# Business Shutdowns and COVID-19 Mortality

**DOI:** 10.1101/2020.10.06.20207910

**Authors:** Gabriele Ciminelli, Sílvia Garcia-Mandicó

## Abstract

Governments around the world have adopted unprecedented policies to deal with COVID-19. This paper zooms in on business shutdowns and investigates their effectiveness in reducing mortality. We leverage upon highly granular death registry data for over 4,000 Italian municipalities in a diff-in-diff approach that allows us to credibly mitigate endogeneity concerns. Our results, which are robust to controlling for a host of co-factors, offer strong evidence that business shutdowns are very effective in reducing mortality. We calculate that the death toll from the first wave of COVID-19 in Italy would have been twice as high in their absence. Our findings also highlight that timeliness is key – by acting one week earlier, the government could have reduced the death toll by an additional 25%. Finally, our estimates suggest that shutdowns should be targeted: closing shops, bars and restaurants saves the most lives, while shutting down manufacturing and construction activities has only mild effects.

## 1 Introduction

The sudden appearence of COVID-19 in early 2020 has forced many governments around the world to implement hastily arranged policies to limit the death toll from the virus, including travel restrictions, lockdowns, and business shutdowns. These measures aim to save lives by reducing the rate of new infections – “flattening the curve” – thus preventing the congestion of the health care system and ensuring that there is enough capacity to admit everyone in need (Ferguson et al., 2020). In this paper, we investigate the effectiveness of business shutdowns, focusing on three issues in particular. First, we assess the overall effects of business shutdowns in reducing mortality. Second, we focus on the importance of timing – that is, the effect of ordering business shutdowns depending on where we are on the epidemic curve. Third, we explore heterogeneities in the effectiveness of business shutdowns across different sectors of the economy.

To carry out the analysis, we focus on the eight regions of Italy’s north – for long the world’s ground zero for COVID-19. Focusing on Italy’s north has a key advantage in our context: the availability of highly granular daily death registry data, as well as data on a host of socio-demographic, labor market and territorial characteristics, for thousands of municipalities. In particular, our dataset covers more than 4,000 municipalities and spans throughout the entire period of the first wave of the COVID-19 epidemic in Italy (February to May 2020). We use this extraordinary wealth of data, which to our knowledge is unmatched in other countries, in a diff-in-diff approach à la Rajan and Zingales (1998), exploiting a peculiarity in the implementation of the business shutdowns policy that allows us to credibly mitigate endogeneity concerns. Rather than targeting specific firms in specific communities, the government ordered the closure – throughout the country and at the same time – of all firms not operating in essential sectors. By calculating the employment share in non-essential sectors, we are thus able to isolate variation in the degree that communities were affected by the shutdowns. Because the government’s list of essential sector was valid at the national level, such variation is exogenous to the policy itself.

Our analysis offers strong evidence that business shutdowns can be very effective in curbing COVID-19 mortality. Our first finding concerns their overall effectiveness. We estimate that a 10 percentage point higher share of employment in shutdown sectors is associated to 0.7 less deaths per day per 100,000 inhabitants. To put this in context, we use our coefficients to perform some back-of-the-envelope calculations and gauge the number of lives saved through business shutdowns: we calculate that the death toll of the first wave of the COVID-19 epidemic in Italy might have been twice as high in absence of business shutdowns.

Our second set of results highlights instead the importance of timeliness. Exploiting the fact that communities were at different stages in the epidemic curve when business shutdowns were ordered – some had a high number of infections, while others had just a few – we estimate that “early” implementation increases the effectiveness of shutdowns. Had the government ordered the shutdowns one week earlier – when the cumulative case load was about one third lower – it could have prevented an additional 25% of COVID-19 deaths.

In a next step, we show that the effectiveness of business shutdowns depends on which particular sectors are closed down. Our third key finding is that closing down the retail trade and the hospitality sectors has the largest effects in reducing mortality. Instead, shutting down construction and low-tech manufacturing activities – in which interpersonal contact is limited to workers in the same unit – only has mild effects, while the shutdown of the high-tech manufacturing sector – where a high share of work can be done digitally (Manyika et al., 2015), thus minimizing interpersonal contact even more – is completely ineffective.^1^ This heterogeneity in the effectiveness of business shutdowns across sectors is consistent with the notion that a higher degree of interpersonal contact between workers and customers can facilitate the spread of infectious diseases (see Dingel and Neiman, 2020; Markowitz et al., 2019; Lewandowski et al., 2020, among others).

Our results speak to a quickly growing literature that uses epidemiological models (such as SIR and SEIR) to assess how mitigation policies can be optimally set to minimize the burden on the economy while reducing the number of fatalities (see, among many others, Atkeson, 2020; Eichenbaum et al., 2020; Jones et al., 2020). To ensure tractability, SIR and SEIR models typically assume an equal number of contacts for each individual (Khain, 2020).^2^ Our results show that this assumption may be problematic when determining optimal business shutdown policies.

We also contribute to a broader literature quantifying the effectiveness of social distancing policies on COVID-19 mortality. A large part of this literature has focused on evaluating the effects of lockdowns and travel restrictions on contagion and, ultimately, mortality (Adda, 2016; Becchetti et al., 2020; Chinazzi et al., 2020; Fang et al., 2020; Ferguson et al., 2020; Juranek and Zoutman, 2020; Pedersen and Meneghini, 2020). Our focus is instead on the effects of businesses shutdowns. To the best of our knowledge, we are the first to quantify the effectiveness of such policies.

Our results relate to Lewandowski et al. (2020) and Muellbauer and Aron (2020), who explore COVID-19 mortality differences by occupational exposure. The former finds that physical contact through occupational exposure can explain up to a quarter of the spread and mortality of COVID-19 across Europe. The latter find that most of COVID-19 deaths in England were among people employed in the consumer-facing service sector.

The rest of the paper is organized as follows: Section 2 describes the dataset and presents the empirical methodology. Section 3 presents our baseline results on the overall effectiveness of business shutdowns, discusses a battery of robustness checks and illustrates the back-of-the-envelope calculations that we perform to gauge the number of lives saved through the shutdowns. Section 4 extends the analysis to the timing of shutdowns and the heterogeneity of their effectiveness across sectors. Section 5 concludes.

## 2 Dataset and Methodology

### 2.1 Death registry data

Any empirical study on the effectiveness of COVID-19 mitigation and/or suppression policies needs to deal with the issue of how to measure COVID-19 mortality. At first glance, using official fatality data provided by the authorities may appear as the most natural choice. However, as we show in a companion paper (Ciminelli and Garcia-Mandicó, 2020), these data suffer from serious undercounting issues and thus risk understating the true extent of COVID-19 outbreaks. We therefore rely on the concept of excess deaths – that is, the difference between deaths for all causes during the COVID-19 epidemic and deaths that would be expected under normal circumstances. This measure does not allow uncovering the COVID-19 fatality rate, but rather pools together direct and indirect COVID-19 deaths.^3^ Since our focus is not narrowly on the COVID-19 fatality rate, that is not a drawback in our context. Moreover, focusing on excess deaths has the key advantage that underlining data are much more granular than official fatality data, which is crucial for our identification strategy, as it will become clear in Section 2.3 below.

We therefore source death registry data from ISTAT (2020*b*). The data provide municipality-level information on daily deaths for the January/1^st^ to May/15^th^ period, for the years 2015 to 2020 for the majority of Italian municipalities. Our focus is on the eight regions of Italy’s north (Emilia-Romagna, Friuli-Venezia Giulia, Liguria, Lombardy, Piedmont, Trentino-Alto Adige, Valle d’Aosta, and Veneto). These make up for about 50% of Italy’s population and 85% of its fatalities during the first wave of COVID-19. The data cover 4,100 municipalities, together accounting for almost 96% of the local population in the eight regions that we consider. Table A1 in the Appendix provides more descriptive statistics.

Figure 1 below takes a first look at the data. Particularly, it compares daily deaths in 2020 (red solid line) to deaths in the five preceding years. Deaths in 2020 exhibit quite similar trends to deaths in 2016 (blue dashed line), up to the day in which Italy’s first community case was detected (February/21^st^, vertical maroon line). The subsequent increase is exponential. At the their peak, roughly a month after the first detection, daily deaths in 2020 are about two and half times as those in 2016, underscoring the severe effects of COVID-19 on mortality. After the peak, they gradually decline and return to normal levels around the end of our sample (mid-May). Our data thus cover the entire period of the first wave of COVID-19 in Italy.

**Figure 1:**
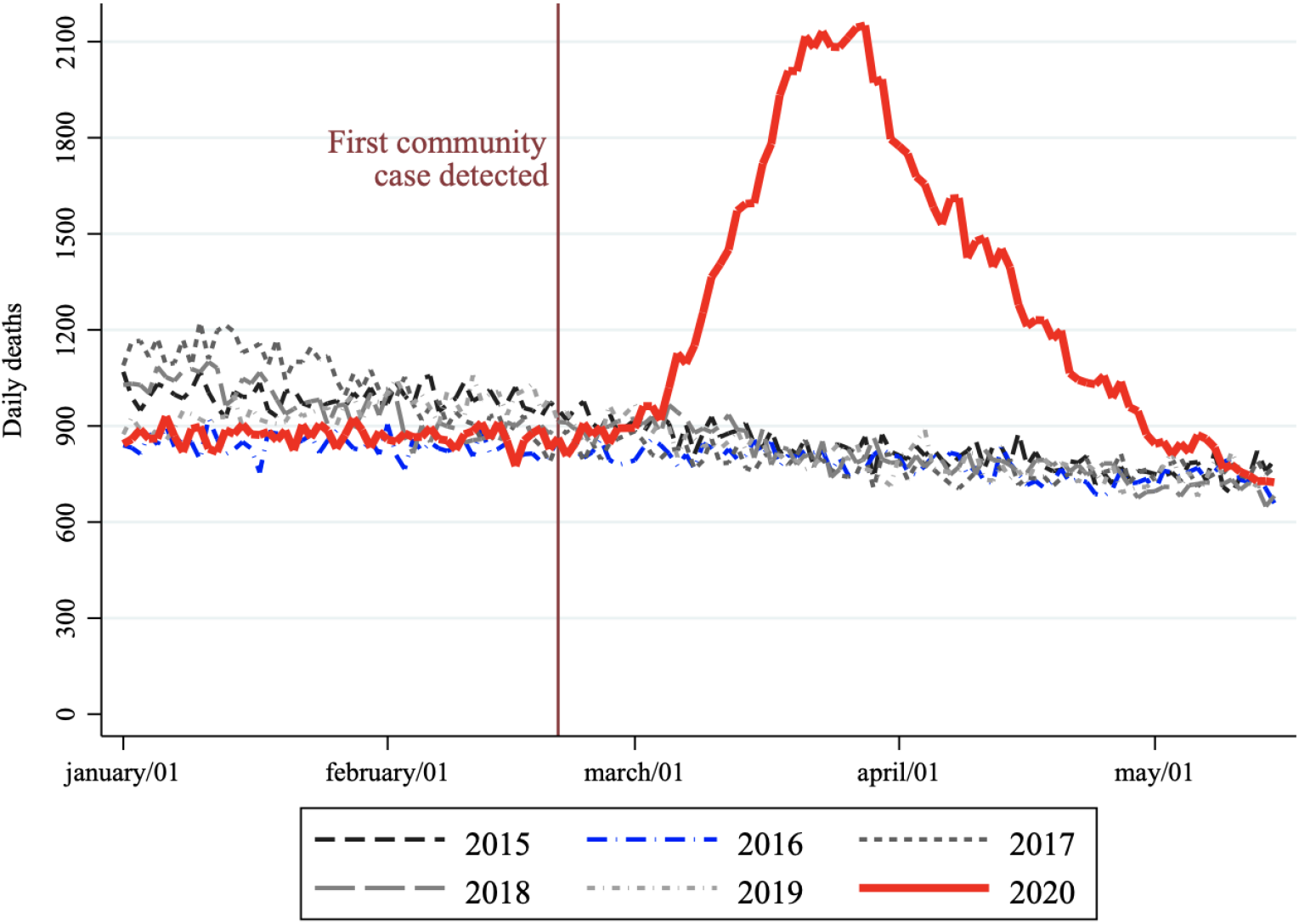
Deaths in 2020 compared to the five previous years. *Notes:* The figure compares daily deaths in 2020 (red solid line) to daily deaths in each of the five preceding years. The blue dashed line denotes deaths in 2016, which we use as counterfactual for the estimation of the effects of COVID-19 on mortality (see Section 2.4). The vertical maroon line denotes the day in which the first COVID-19 community case was detected, on February/21^th^).

### 2.2 Other data

The dataset is complemented with other variables that are needed to carry out the analysis. We start by sourcing census population data (from ISTAT, 2020*c*), which we use to construct the mortality rate series. These data provide information on the resident population in each municipality, as of January/1^st^, for the years 2015 to 2019. We impute 2020 population by using 2019 growth rates. We then turn to business shutdowns. As we discuss more in detail in Section 2.3 below, our empirical approach exploits heterogeneities in the extent that communities are affected by shutdowns. To measure such heterogeneities, we source municipality-level data on the number of workers (both employees and self-employed) in each 3-digit non-farm private sector, from ISTAT (2020*a*).^4^ Since the government also ordered the closure of all educational institutions, we collect data on the number of residents enrolled at university from ISTAT (2020*a*) to run a robustness check.

Finally, we source variables capturing slow-moving municipality characteristics that we use to control for potential co-factors of COVID-19 mortality. These characteristics broadly fall in three categories: (i) labor market characteristics, including the level of internal commuting and the employment rate; (ii) territorial characteristics, such as population density and the level of air pollution; and (iii) socio-demographic characteristics, including the share of females and that of high school graduates in the working age population, as well as the share of the elderly and mean income. We also construct a digital labor index measuring how much work is/can be done remotely in firms that are not affected by the shutdowns.^5^ As most of these variables are not available at a regular frequency, we compute their means over the 2015-2019 period and treat them as time-invariant factors. Table A2 in the Appendix provide information on sources and coverage.

### 2.3 Construction of the business shutdowns variable

The Italian government took unprecedented measures to fight the first wave of the COVID-19 epidemic in 2020. Twelve days after the first community case was identified, it ordered all educational institutions to close down and switch to online learning (March/5^th^). Less than a week later, it introduced a nationwide lockdown and ordered the closure of all non-essential commercial activities, such as shops, bars and restaurants (March/11^th^). About 10 days later, it compounded those measures by shutting down all non-essential production activities (March/22^nd^).^6^ As the situation gradually improved, the government lifted the lockdown and allowed most businesses to reopen after almost two months of restrictions (May/4^th^).

Our focus is on business shutdowns. Did they contribute to reduce mortality? If so, by how much? To answer these questions, we follow the approach of Rajan and Zingales (1998) and exploit variation across communities in the employment share in firms that were forced to shut down, which, crucially, is exogenous to the government policies. Exogeneity follows from the fact that, rather than targeting specific firms in specific communities, the government uniformly ordered the closure of all firms in certain specific sectors throughout the country. To do so, it issued a list of essential sectors valid countrywide: firms operating in those sectors could stay open, while all the others had to close down. Our identifying assumption is that business shutdowns were more binding in labor markets with a higher employment share in non-essential sectors (those in which firms had to shut down).

We therefore construct a variable measuring the employment share in firms affected by the shutdowns by matching the government’s list of essential sectors to data on the number of workers in each non-farm private sector, sourced from ISTAT (2020*a*). Following common practice in spatial and urban economics studies (see Moretti, 2010; Acemoglu and Restrepo, 2020; Allcott and Keniston, 2018; David et al., 2013, and others), we pool municipalities into local labor markets. Essentially, these are areas whose borders are defined in a way that minimize external commuting (commuting outside borders). Since in Italy many people work or study in a different municipality than the one in which they reside, carrying out the analysis at the local-labor-market-level limits possible mismatches between the extent to which individuals are affected by the government policies and the municipality where COVID-19 deaths are recorded. Overall, we follow 222 local labor markets, each with a population of about 125,000 inhabitants on average.^7^ Next, we calculate the number of workers in each sector affected by the government policies and construct our business shutdowns variable, *BS*_*it*_, according to the following formula:^8^

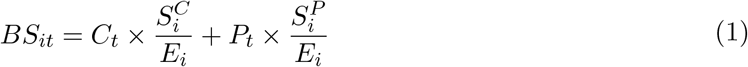

where 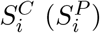 measures the number of workers in commercial (production) firms affected by the shutdown of non-essential activities; *E*_*i*_ is total non-farm private employment; and *C*_*t*_ (*P*_*t*_) is a commercial (production) activities shutdown policy dummy that takes value equal to 1 from 7 days after the introduction of the policy to 7 days after its expiration (the 7-day lag is in order to account for the time elapsing between COVID-19 infection and death, see Lauer et al., 2020; Linton et al., 2020; SARS-CoV-2 Surveillance Group, 2020, among others).^9^

Our business shutdowns variable recognizes that commercial and production activities were affected at different times and thus it is time-varying. When both non-essential commercial and production activities were shut, about 50% of all non-farm private sector workers were forced to stay home, on average. Importantly, there is large variation across local labor markets, with the standard deviation of the employment share in shutdown firms being more than 9%.

### 2.4 Empirical specifications

In this section, we describe our empirical methodology. We start by illustrating how we estimate the unconditional effects of COVID-19 on mortality, while we discuss how we investigate the effectiveness of business shutdown policies in reducing mortality further below. For the estimation, we opt for a differences-in-differences approach, using the year 2016 as counterfactual of what mortality would have been in absence of COVID-19. This gives us a measure of the excess deaths from COVID-19.^10^ We normalize the within-year time dimension *t* to take value equal to 0 on the day in which the first community case was detected (February/21^st^) and negative (positive) values on days before (after). We then estimate the mortality effects of COVID-19 over time through a flexible specification, as follows:

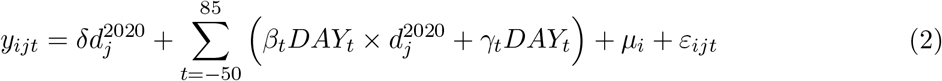

where *y*_*ijt*_ measures daily deaths per 100,000 inhabitants in local labor market *i*, at within-year time *t*, for year *j*; 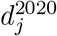 is a dummy variable taking value equal to 1 in 2020 and 0 otherwise; *DAY*_*t*_ are within-year time effects, taking value 1 in each particular day of the year and 0 otherwise; *µ*_*i*_ are local labor markets fixed effects; and *ε*_*ijt*_ is an idiosyncratic error, clustered at the level of local labor markets. The coefficients of interest are the *β*_*t*_s, which capture the effect of COVID-19 on the mortality rate at time *t*. For the estimation, we use the least squares method with population analytical weights.^11^

Figure A2 in the Appendix shows the effect of COVID-19 on daily deaths per 100,000 inhabitants, as estimated through Equation (2). The Figure confirms the validity of the parallel trends assumption, and shows that the effect of COVID-19 on mortality peaks 33 days after onset, at about five deaths per day per 100,000 inhabitants, to then slowly decrease, until becoming statistically insignificant 39 days after the peak. These estimates are robust to using average deaths in the five preceding years (2015 to 2019) as counterfactual (also reported in Figure A2).

We next turn to business shutdowns. To estimate their effects on COVID-19 mortality, we extend Equation (2) by adding an interaction between the year 2020 dummy 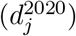 and the variable measuring the employment share in shutdown activities (*BS*_*it*_). The specification that we estimate is as follows:

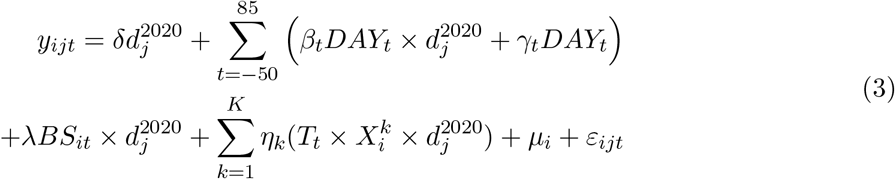

where *T*_*t*_ is a COVID-19 treatment dummy, taking value equal to 1 in the period after the detection of the first community case and 0 in the period before; the 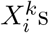 are (time-invariant) local labor market characteristics included as controls; and the rest is as in Equation (1). The *λ* coefficient measures the effect of having a higher employment share in shutdown firms on the COVID-19 mortality rate. As before, we estimate Equation (3) through OLS with analytical population weights and standard errors clustered at the level of the local labor market.

## 3 The Effects of Business Shutdowns on Mortality

### 3.1 Main Results

Table 1 below reports the results on the effect of business shutdowns in reducing COVID-19 mortality. Column (1) shows estimates from a parsimonious specification with only the share of females in the working age population as control, which we include since (i) gender is an important factor in determining COVID-19 mortality (SARS-CoV-2 Surveillance Group, 2020) and (ii) females constitute the vast majority of workers in the health care and other essential sectors at high risk of COVID-19 exposure (Boniol et al., 2019). Columns (2)-(5) show estimates from extended specifications in which we gradually add other demographic factors and labor market as well as territorial characteristics as additional controls. The reported coefficients measure the changes in daily deaths per 100,000 inhabitants associated with a standard deviation increase in the variable considered (9% in the case of the business shutdowns variable).

**Table 1:** The effects of business shutdowns on COVID-19 mortality.

|  | (1) | (2) | (3) | (4) | (5) |
| --- | --- | --- | --- | --- | --- |
| Business shutdowns | -0.67***<br>(0.13) | -0.82***<br>(0.18) | -0.62***<br>(0.14) | -0.61***<br>(0.14) | -0.69***<br>(0.14) |
| Share working age females | -0.98***<br>(0.21) | -0.71***<br>(0.20) | -0.67***<br>(0.22) | -0.67***<br>(0.22) | -0.70***<br>(0.21) |
| Share high school graduates |  | -0.37*<br>(0.20) | -0.58**<br>(0.24) | -0.57**<br>(0.25) | -0.49**<br>(0.25) |
| Population density |  |  | 0.28***<br>(0.09) | 0.27***<br>(0.10) | 0.34***<br>(0.09) |
| Days PM10 above limit |  |  |  | 0.03<br>(0.11) | 0.18<br>(0.12) |
| Digital labor in active firms |  |  |  |  | -0.45***<br>(0.16) |
| Observations | 60,384 | 60,384 | 60,384 | 60,384 | 60,384 |
| R-squared | 0.233 | 0.235 | 0.242 | 0.242 | 0.245 |

**Notes:** The first row in this Table reports the effects of having a one-standard deviation higher employment share in non-essential activities. This is measured by the $\lambda$ coefficient from Equation (3). Column (1) includes only the share of working age females as control variable, while columns (2)-(5) expand the specification to incorporate other control variables. Reported coefficients of control variables are normalized to measure the effect of a one standard deviation increase in the variable. The dependent variable is daily deaths per 100,000 inhabitants. All specifications include local labor market fixed effects, and are obtained using least squares with analytical population weights. Standard errors are clustered at the local labor market level.
Significance levels: \* $p \leq 0.1$ , \*\* $p \leq 0.05$ , \*\*\* $p \leq 0.01$ .

We estimate a negative and highly statistically significant coefficient for our business shutdowns variable across all specifications. A standard deviation higher employment share in shutdown firms is associated with 0.6 to 0.8 less deaths per day per 100,000 inhabitants, depending on the specification considered. The coefficient is −0.7 in our most comprehensive specification including all controls, which we use as baseline in the rest of the analysis (Column (5)). Compared with an average daily death toll of about 1.8 deaths per 100,000 inhabitants, these results indicate that business shutdowns have quite meaningful effects in reducing mortality. We better quantify these effects in Section 3.2 below, where we perform some back-of-the-envelope calculations on the overall number of deaths that have been prevented through these policies.

Turning to the control variables, they all enter with the expected sign and, except in one case, they are also highly statistically significant. A higher share of women in the working age population is by far the single most important factor reducing COVID-19 mortality. We also find an important mitigating role for education, proxied by the share of high school graduates.^12^ Among risk factors, we control for population density and the level of air pollution. We estimate a positive and statistically significant coefficient for the former, confirming that closer interpersonal proximity facilitates the spread of COVID-19, while our results for the latter are inconclusive.^13^ Finally, our digital labor index measuring how much work can be done remotely in firms that stay open has a negative and highly significant coefficient, indicating that remote working is associated with lower mortality.

#### Robustness Checks

Table A3 in the Appendix reports estimates from a battery of robustness checks on our baseline estimates. For ease of reference, Column (1) reports estimates from our baseline specification including all controls (as in Column (5) of Table 1 above). We start by verifying that our results do not depend on the choice made to account for the lag between the timing of the policy implementation and its effects on mortality (which is 7 days in our baseline) and estimate two alternative specifications in which (i) we increase this lag to 14 days, and (ii) do not not account for a lag at all. The new results are very similar to, and not statistically different from, our baseline (Columns (2) and (3)).

Next, we focus on another aspect of the construction of the business shutdowns variable. Although the government never formally ordered the shutdown of hotels, the accommodation sector was effectively shut since the introduction of the countrywide lockdown, as this severely restricted the movement of people and brought tourism to a halt. For this reason, in constructing the business shutdowns variable used in our baseline we also included the accommodation sector among shutdown commercial activities (the 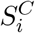 component in Equation (1) above). We estimate a specification in which we exclude the accommodation sector from the 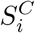 variable. Again, the results are very similar to, and not statistically different from, our baseline (Column (4)).

We also run two placebo tests to verify that our estimates are not the result of spurious relationships. In the first, we replace our policy intervention dummies *C*_*t*_ and *P*_*t*_ in Equation (1) with dummies taking value equal to 1 in the pre-policy period and equal to 0 otherwise. In the second, we instead assign random values to the 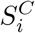 and 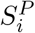 variables in Equation (1). Reassuringly, the new estimated coefficients are not statistically significant, suggesting that our baseline estimates are not spurious (Columns (5) and (6)). Finally, we also check that our results do not suffer from omitted variable bias and add the employment rate, an index measuring the level of internal commuting, the share of residents enrolled at university, the share of people older than 80 and the mean income as additional controls.^14^ The new estimates on the effects of business shutdowns are very close to, and not statistically different from, our baseline (Column (7)). All in all, we conclude that our baseline results are robust.

### 3.2 Back-of-the-envelope calculations on saved lives

How many deaths can be ascribed to COVID-19? And how many deaths have been prevented by shutting businesses down? We answer these questions through some simple back-of-the-envelope calculations. We start by using the coefficients estimated through Equation (3) to calculate the number of deaths caused by COVID-19, in each day of the epidemic and each local labor market, according to the following formula:

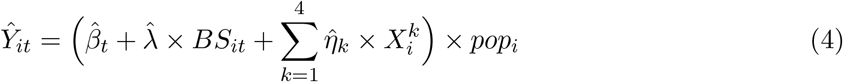

where *Ŷ*_*it*_ is the predicted number of deaths due to COVID-19 in local labor market *i* in day *t*; the 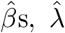, and 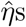 are the coefficients estimated from Equation (3); *BS*_*it*_ is the employment share in shutdown firms; the *X*s are the control variables discussed above; *pop*_*i*_ is population (in 100,000). We calculate *Ŷ*_*it*_ using the coefficients reported in Column (5) of Table 1 and then sum each *Ŷ*_*it*_ over the entire period of the first wave of the COVID-19 epidemic in Italy (February/21^st^ to May/15^th^ period), and across all local labor markets. This simple back-of-the-envelope calculation indicates that COVID-19 contributed to the death of almost 45,000 people – about 0.15% of the local population – during the first wave of the epidemic in the eight regions of Italy’s north.

Next, to get a rough sense of the efficacy of the business shutdown policies in reducing mortality, we calculate the number of deaths that would have resulted in a hypothetical scenario of no business shutdowns. To do so, we artificially set the variable *BS*_*it*_ in Equation (4) equal to 0 and find that deaths would have been almost 90,000 – about twice as much as those that actually occurred – in absence of business shutdowns. Our estimates thus suggest that business shutdowns halved COVID-19 deaths. Two caveats are however in order. First, in Italy the business shutdowns coincided with the lockdown period. Hence, our results are conditional on a lockdown being in place and should not be generalized to cases in which business shutdowns are disjoint from lockdown policies. Second, it may be that some non-essential businesses would have closed even in the absence of government-mandated policies. If this were to be the case, we would be overestimating the deaths that were saved through the government policies.

## 4 Extensions

### 4.1 The importance of timely interventions

Given that infections grow exponentially, the effects of mitigation and suppression policies should be stronger if implemented earlier in the epidemic curve. For instance, Dave et al. (forthcoming) and Friedson et al. (2020) show that this is indeed true for the case of lockdowns. In this section, we investigate to which extent this is valid for business shutdowns. We exploit the fact that, while the policies adopted by the government were applied at the same time throughout the entire country, communities were at different stages in their epidemic curve when they were affected by the shutdowns.^15^ Hence, the variation across local labor markets in the timing of the policy implementation relative to the stage of the epidemic can be considered as being exogenous to the policy itself.

We proceed by constructing a variable measuring differences across local labor markets in the relative “timing” of the policy implementation. To do so, we take the ratio of the cumulative number of cases reported up to the day in which the government policies were implemented (sourced from Protezione Civile, 2020) to the population. On average, detected cases were about 1,400 per million people when the business shutdowns were adopted. But there was great heterogeneity across local labor markets. Some had recorded as little as 70 cases per million people, while others had almost 17,000. To estimate the importance of timing, we augment Equation (3) by adding an interaction term that is the product of the timing variable, the variable measuring the employment share in shutdown sectors (*BS*_*it*_) and the year 2020 dummy 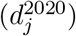.^16^

Table 2 below shows the results. For ease of reference, Column (1) reports the results from our baseline specification (as in Column (5) of Table 1 above). Column (2) shows estimates from the extended specification accounting for the timing of the business shutdowns relative to the stage of the epidemic. The interaction between the business shutdowns and the timing variables enters with a positive sign and it is highly statistically significant, providing evidence that communities in which the policy was implemented at a later stage in the infection curve benefited less from it. Specifically, the effectiveness of business shutdowns in reducing mortality is about 15% lower in local labor markets with a cumulative case load at the time of implementation one standard deviation higher than average. In the extreme cases of local labor markets in which COVID-19 was at its most advanced stage, the effectiveness of business shutdowns is reduced by almost 70%.

**Table 2:**
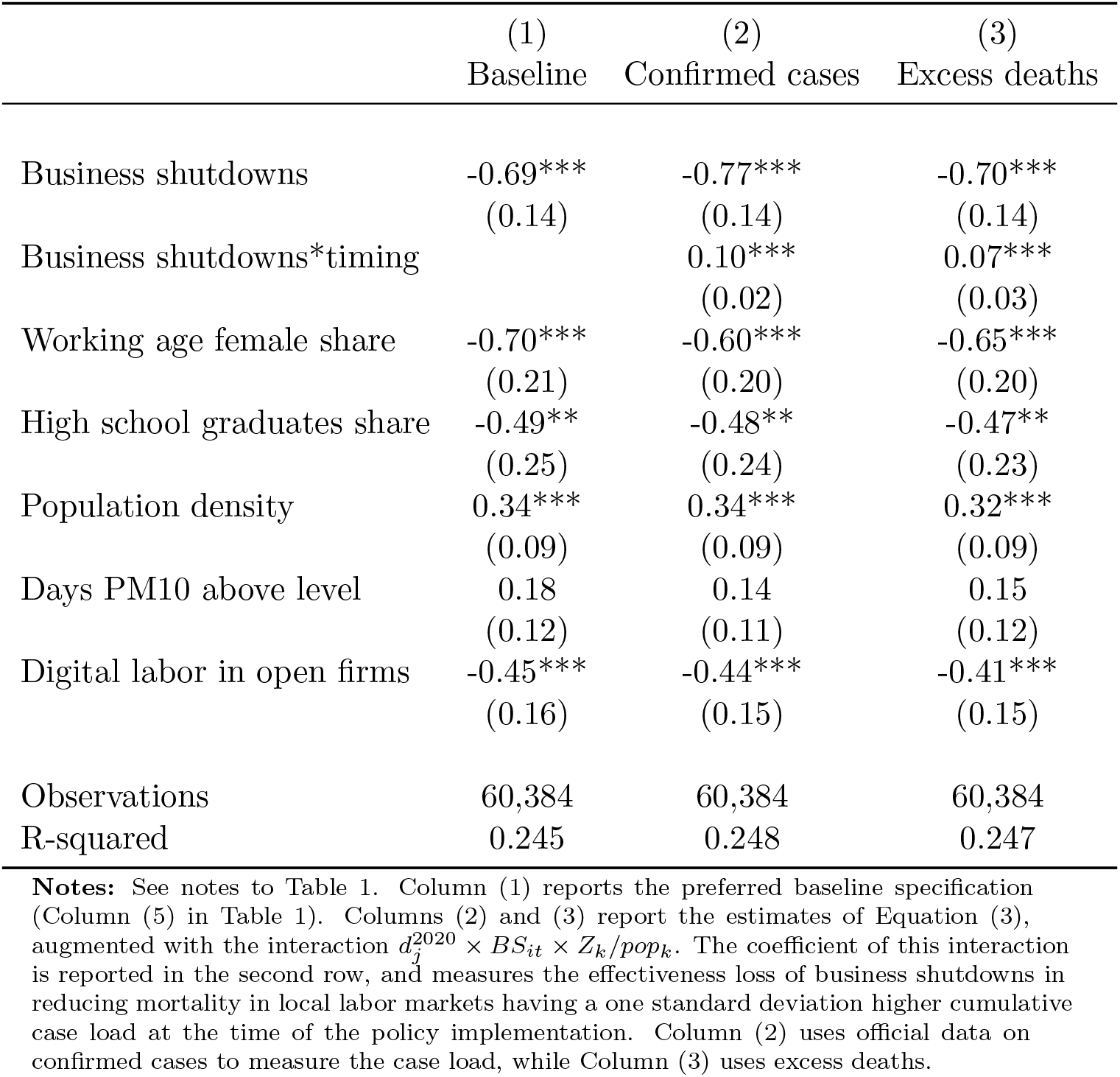
The effects of business shutdowns accounting for timing.

|  | (1)<br>Baseline | (2)<br>Confirmed cases | (3)<br>Excess deaths |
| --- | --- | --- | --- |
| Business shutdowns | -0.69***<br>(0.14) | -0.77***<br>(0.14) | -0.70***<br>(0.14) |
| Business shutdowns*timing |  | 0.10***<br>(0.02) | 0.07***<br>(0.03) |
| Working age female share | -0.70***<br>(0.21) | -0.60***<br>(0.20) | -0.65***<br>(0.20) |
| High school graduates share | -0.49**<br>(0.25) | -0.48**<br>(0.24) | -0.47**<br>(0.23) |
| Population density | 0.34***<br>(0.09) | 0.34***<br>(0.09) | 0.32***<br>(0.09) |
| Days PM10 above level | 0.18<br>(0.12) | 0.14<br>(0.11) | 0.15<br>(0.12) |
| Digital labor in open firms | -0.45***<br>(0.16) | -0.44***<br>(0.15) | -0.41***<br>(0.15) |
| Observations | 60,384 | 60,384 | 60,384 |
| R-squared | 0.245 | 0.248 | 0.247 |

**Notes:** See notes to Table 1. Column (1) reports the preferred baseline specification (Column (5) in Table 1). Columns (2) and (3) report the estimates of Equation (3), augmented with the interaction $d_j^{2020} \times BS_{it} \times Z_k/pop_k$ . The coefficient of this interaction is reported in the second row, and measures the effectiveness loss of business shutdowns in reducing mortality in local labor markets having a one standard deviation higher cumulative case load at the time of the policy implementation. Column (2) uses official data on confirmed cases to measure the case load, while Column (3) uses excess deaths.

In carrying out this analysis, we measured the relative timing of the intervention by using official data on confirmed COVID-19 cases. That is our preferred approach, since these data are readily available to policymakers when they have to decide on new policies. As long as there is not a systematic pattern in the undercounting of COVID-19 cases across local labor markets (see discussion in Section 2.1), the fact that official data are likely to understate the true extent of the epidemic is not an issue for the purpose of measuring variation across communities in the relative timing of the policy implementation. However, we also estimate an alternative specification in which we use data on excess deaths in 2020 relative to 2016, rather than official data on detected cases, to measure the relative implementation timing. We report results from this alternative specification in Column (3) of Table 2. The new estimates are very close to, and not statistically different from, those obtained through our preferred approach, which uses official case data.

Our results beg the natural question of how many more lives could have been saved had the government intervened earlier in the epidemic curve. To answer this question we use our estimates from Column (2) of Table 2 above to perform some back-of-the-envelope calculations, similarly to what done in Section 3.2. We find that the death toll from COVID-19 would have been about 33,000 – more than 25% lower – had the business shutdowns been implemented one week earlier, when cumulative cases cases per million inhabitants were around 500 on average.^17^

### 4.2 Heterogeneity in the effectiveness of business shutdowns across sectors

The analysis carried out so far suggests that business shutdowns are generally quite effective in reducing COVID-19 mortality. However, there may exist heterogeneities depending on which sectors are affected. Consumer-facing service activities have potential to amplify contagion due to the high degree of interpersonal contact between workers and customers (Markowitz et al., 2019; Lewandowski et al., 2020), while workers in industries where a high share of work can be done remotely should be more shielded by contagion. This raises the natural question of whether the effectiveness of business shutdowns depends on the particular sectors that are closed down. Answering this question is of crucial importance: if the answer was positive, policy-makers could implement targeted interventions that maximize lives saved and minimize the economic costs.

In this section, we extend the analysis to explore such heterogeneties. We start by dividing the economy in twenty broad sectors, following the ISIC Rev 4 industry classification, and consider the different extent through which each sector was affected by the shutdowns.^18^ Table 3 below reports relevant sector-specific statistics, such as the share in overall non-farm private employment, the employment share in shutdown firms and the propensity towards digital labor (from Manyika et al., 2015), which may be a factor driving heterogeneity in the effectiveness of business shutdowns. Looking at the share in overall employment and the employment share in shutdown firms, we notice that the five largest sectors – low-tech manufacturing, hospitality, retail trade, construction and high-tech manufacturing – are also among those most impacted by the shutdowns: together they account for about 60% of all employment, but they make up for almost 80% of employment in shutdown firms. As shown by Navaretti et al. (2020), they are also the most systemically important for the economy.^19^ For these reasons, in the analysis that follows we zoom in on these five sectors. Looking at the propensity towards digital labor, high-tech manufacturing has the second highest score, while retail trade and hospitality receive the lowest; construction and low-tech manufacturing are in the middle.

**Table 3:** Business shutdowns by sector.

|  | industry<br>code | empl.<br>share | shutdown<br>YES/NO | share<br>mean | affected<br>s.d. | digital<br>labor |
| --- | --- | --- | --- | --- | --- | --- |
| Mining and Quarrying | B | 0.11 | YES | 99.83 | 1.02 | 1 |
| High-tech manufacturing | C* | 8.62 | YES | 67.42 | 22.32 | 5 |
| Low-tech manufacturing | C** | 19.27 | YES | 55.27 | 17.00 | 3 |
| Electricity and Gas | D | 0.50 | NO | / | / | 4 |
| Water/waste management | E | 0.83 | NO | / | / | / |
| Construction | F | 9.43 | YES | 63.64 | 7.85 | 2 |
| Sales of motorveichles | 45 | 2.09 | YES | 19.64 | 10.73 | / |
| Wholesale trade | 46 | 5.49 | YES | 64.14 | 10.64 | 4 |
| Retail trade | 47 | 10.75 | YES | 40.78 | 3.92 | 1 |
| Transportation & storage | H | 4.93 | NO | / | / | 2 |
| Hospitality | I | 11.88 | YES | 94.47 | 4.44 | 1 |
| Information & communication | J | 1.46 | NO | / | / | 6 |
| Finance/insurance activities | K | 0.83 | NO | / | / | 5 |
| Real estate activities | L | 2.09 | YES | 100.00 | 0.00 | 2 |
| Professional activities | M | 5.50 | YES | 2.59 | 1.73 | 5 |
| Administrative activities | N | 4.62 | YES | 19.46 | 14.31 | 5 |
| Education | P | 0.59 | NO | / | / | 3 |
| Health | Q | 4.17 | NO | / | / | 2 |
| Entertainment & recreation | R | 0.93 | YES | 100.00 | 0.00 | 1 |
| Other service activities | S | 2.60 | YES | 81.88 | 0.99 | 3 |

**Notes:** This table divides non-farm private employment in twenty broad sectors. The first and second columns report the name and code, according to the ISIC Rev 4 classification, of each sector. The third column states whether a sector is affected by the shutdowns or not. The fourth and fifth columns report the mean and standard deviation of the employment share in shutdown firms. The sixth column reports the score from the digital labor index of Manyika et al. (2015). High-tech manufacturing includes the 20, 21, 254, 26, 27, 28, 29, 30 and 325 industries. Low-tech manufacturing includes all industries between 10 and 33, except those belonging in high-tech manufacturing.

For the estimation, we twist Equation (3) and replace the variable *BS*_*it*_ with sector-specific variables measuring, within each sector, the employment share in firms that are closed down. Table 4 below reports the results. In Columns (1)-(5) we show estimates when considering one sector at a time, while Column (6) reports estimates from a comprehensive specification including all five sectors. The coefficients report the change in daily deaths per 100,000 inhabitants associated with a 10 percentage points increase in the employment share in shutdown firms. All specifications also include our baseline control variables (as in Column (5) of Table 1). For the sake of brevity, we only report the coefficient of the digital labor variable, which is the main variable of interest (all other controls display coefficients that are very similar to those reported in Table 1).

**Table 4:** Sector-specific effects of business shutdowns.

|  | (1) | (2) | (3) | (4) | (5) | (6) |
| --- | --- | --- | --- | --- | --- | --- |
| Retail trade | -0.77**<br>(0.36) |  |  |  |  | -0.71**<br>(0.34) |
| Hospitality |  | -0.71**<br>(0.33) |  |  |  | -0.66*<br>(0.36) |
| Construction |  |  | -0.41**<br>(0.17) |  |  | -0.25<br>(0.17) |
| Low-tech manufacturing |  |  |  | -0.11<br>(0.08) |  | -0.17**<br>(0.08) |
| High-tech manufacturing |  |  |  |  | -0.03<br>(0.05) | -0.01<br>(0.05) |
| Digital labor in active firms | -0.33**<br>(0.15) | -0.44***<br>(0.16) | -0.34**<br>(0.15) | -0.28*<br>(0.15) | -0.32**<br>(0.15) | -0.40***<br>(0.15) |
| Observations | 59,568 | 59,568 | 59,568 | 59,568 | 59,568 | 59,568 |
| R-squared | 0.241 | 0.242 | 0.241 | 0.240 | 0.240 | 0.244 |

**Notes:** See notes to Table 1. This table reports results for the sector-specific effects of business shutdowns in reducing mortality. Columns (1)-(5) report coefficients when each sector is considered one at a time. Column (6) considers all sectors at the same time. The coefficients are estimated using the employment share, within each particular sector, in shutdown firms in place of the variable $BS_{it}$ in Equation (3).

Our results point to substantial heterogeneities in the effectiveness of business shutdowns across sectors. Closing down retail trade and hospitality activities is the most effective way to reduce mortality. Shutting down construction and low-tech manufacturing has relatively mild, and not precisely estimated, effects in terms of reduced mortality, while the closure of the high-tech manufacturing sector is not effective at all.

The result that shutting down production activities, such as construction sites and factories, may be less effective in reducing mortality than closing down services is in line with the findings on occupational exposure to COVID-19 of Lewandowski et al. (2020) and Muellbauer and Aron (2020). In particular, the latter uses excess mortality data for England to find that most of COVID-19 deaths in the working-age population were concentrated among people employed in the consumer-facing service sector. While workers in the service sector interact with consumers every day – the opposite of social distancing – for the most part, factory workers only interact with other workers in the same unit, and the opportunities to contract or spread the virus on the workplace appear to be more limited than in the consumer-facing service sector. Our results also have clear policy implications. Governments should not hesitate in closing down services if they want to reduce COVID-19 mortality. On the other hand, they should carefully weight the less clear benefits of closing down factories against the undoubted costs of the halt in production.

## 5 Conclusion

The sudden appearance of COVID-19 in early 2020 has brought about significant human losses in most countries around the world, often forcing governments to implement hastily arranged policies to limit the death toll from the virus. The closure of non-essential commercial and production activities (commonly referred to as business shutdowns) has been one such policy. This paper investigated its effectiveness in reducing COVID-19 mortality by using highly granular daily death registry and employment data for thousands of municipalities in Italy’s north, for long the world’s worst affected area by COVID-19. To carry out the analysis, we exploited a peculiarity in the implementation of the policy in a diff-in-diff approach à la Rajan and Zingales (1998) that allowed us to credibly mitigate endogeneity concerns. Rather than targeting specific firms in specific communities, the government ordered the closure, throughout the country and at the same time, of all firms not operating in essential sectors. By calculating the employment share in shutdown sectors, we were thus able to isolate variation in the degree that communities were affected by the business shutdowns.

Our results, which are robust to controlling for a host of co-factors, offer strong evidence that business shutdowns can be very effective in curbing COVID-19 mortality. Using our estimates to perform some back-of-the-envelope calculations, we calculated that business shutdowns might have halved the death toll of COVID-19 during the first wave of the epidemic in Italy. We also found that implementing business shutdowns early on in the epidemic curve increases their effectiveness. We calculated that if the government had introduced the policy one week earlier it could have reduced the death toll by an additional 25%. We also explored heterogeneity in the effectiveness of business shutdowns across sectors. Consistent with the notion that sectors with a higher degree of interpersonal contact can facilitate the spread of infectious diseases, we found that closing down the retail trade and hospitality sectors has the largest effects in reducing mortality. Shutting down the construction and the manufacturing sectors – in which interpersonal contact is limited to workers in the same unit – only has mild effects.

Our analysis carries clear policy implications. Governments should not hesitate to act early on to order the closure of non-essential businesses to save lives when new outbreaks of COVID-19 and other pathogens materialize. However, since business shutdowns carry large economic costs, they should consider targeted interventions, prioritizing the closure of sectors with a high degree of interpersonal contact between workers and customers and a low propensity toward digital labor.

## Data Availability

Data available upon request

## Appendix

**Table A1:** Population coverage of death registry data.

| Region | mun. | % of all mun. | pop. (in 1,000) | % of all pop. |
| --- | --- | --- | --- | --- |
| Emilia-Romagna | 304 | 92.68 | 4412 | 96.29 |
| Friuli Venezia Giulia | 200 | 93.46 | 1206 | 93.15 |
| Liguria | 218 | 93.56 | 1581 | 97.06 |
| Lombardy | 1455 | 97.00 | 10499 | 98.43 |
| Piedmont | 1099 | 93.69 | 4486 | 95.33 |
| Trentino-Alto Adige | 256 | 88.58 | 1095 | 92.36 |
| Valle d'Aosta | 68 | 91.89 | 134 | 90.88 |
| Veneto | 500 | 87.87 | 4684 | 91.01 |
| Total | 4100 | 93.58 | 28097 | 95.71 |

**Notes:** the columns “mun.” and “% of all mun.” report the number of municipalities covered and their share over all municipalities in the region. The columns “pop (in 1,000)” and “% of all pop.” report the population covered (in thousands) and its share over all the regional population.

**Table A2:** Variable descriptives, source and definitions.

| Variable | obs | mean | s.d. | min | max | Period | Source | Variable definition |
| --- | --- | --- | --- | --- | --- | --- | --- | --- |
| Business shut-downs | 222 | 50.34 | 9.12 | 27.92 | 78.72 | 2017 | ISTAT (2020a) and own calculations | Employment share in shut-down firms (in %) |
| Share working age females | 222 | 49.56 | 0.8 | 46.39 | 51.48 | 2020 | ISTAT (2020c) | Share of women in 18-65 population (in %) |
| Share high school graduates | 222 | 57.88 | 6.14 | 33.34 | 71 | 2015 | ISTAT (2018) | Share of high school graduates in 25-64 population (in %) |
| Population density | 222 | 2442.93 | 779.29 | 927.48 | 6346.89 | 2017 | (ISTAT, 2020d) | Population over inhabited territory land |
| Days PM10 above limit | 222 | 45.05 | 26.03 | 2.33 | 90 | 2015-2016 | (ISTAT, 2019) | Days in a year in which PM10 averages above 50 $\mu\text{g}/\text{m}^3$ (provincial level) |
| Digital labor in active firms | 222 | 0.47 | 0.06 | 0.24 | 0.68 | 2017 | ISTAT (2020a) and own calculations using Manyika et al. (2015) | Weighted sum of employment across industries using digital labor scores as a share of total employment (0-1) |
| University closure | 222 | 26.17 | 6.33 | 2.19 | 41.11 | 2017 | ISTAT (2020d) and own calculations | Share of people enrolled at university over 18-25 population (in %) |
| Internal commuting index | 222 | 36.87 | 12.66 | 7.13 | 66.11 | 2015 | ISTAT (2011) | Flows across municipalities over total flows |
| Share of 80+ | 222 | 7.51 | 1.43 | 4.57 | 12.68 | 2015-2019 | ISTAT (2020c) | People 80+ over total population (in %) |
| Mean income | 222 | 4872.05 | 530.45 | 2656.72 | 6362.4 | 2015-2017 | ISTAT (2020d), ISTAT (2020c) and own calculations | Total income divided by population |

**Table A3:** Robustness checks on the effects of business shutdowns on COVID-19 mortality.

|  | (1)<br>Baseline | (2)<br>Timing<br>14-day | (3)<br>Timing<br>no lag | (4)<br>Sectors<br>affected | (5)<br>Placebo<br>time | (6)<br>Placebo<br>share | (7)<br>Additional<br>controls |
| --- | --- | --- | --- | --- | --- | --- | --- |
| Business shutdowns | -0.69***<br>(0.14) | -0.59***<br>(0.13) | -0.61***<br>(0.15) | -0.62***<br>(0.15) | 0.12<br>(0.19) | 0.04<br>(0.66) | -0.63***<br>(0.15) |
| Share working age females | -0.70***<br>(0.21) | -0.70***<br>(0.21) | -0.70***<br>(0.21) | -0.70***<br>(0.22) | -0.69***<br>(0.22) | -0.70***<br>(0.22) | -0.60***<br>(0.20) |
| Share high school graduates | -0.49**<br>(0.25) | -0.47*<br>(0.25) | -0.48*<br>(0.25) | -0.50*<br>(0.26) | -0.35<br>(0.23) | -0.37<br>(0.25) | -0.69***<br>(0.24) |
| Population density | 0.34***<br>(0.09) | 0.35***<br>(0.10) | 0.34***<br>(0.09) | 0.32***<br>(0.10) | 0.40***<br>(0.11) | 0.39***<br>(0.11) | 0.26**<br>(0.13) |
| Days PM10 above limit | 0.18<br>(0.12) | 0.18<br>(0.12) | 0.18<br>(0.12) | 0.17<br>(0.13) | 0.16<br>(0.12) | 0.16<br>(0.12) | 0.18<br>(0.13) |
| Digital labor in active firms | -0.45***<br>(0.16) | -0.43***<br>(0.16) | -0.44***<br>(0.16) | -0.31**<br>(0.16) | -0.33**<br>(0.15) | -0.32**<br>(0.15) | -0.51**<br>(0.20) |
| University closures |  |  |  |  |  |  | -0.01<br>(0.02) |
| Employment rate |  |  |  |  |  |  | -0.17<br>(0.23) |
| Internal commuting index |  |  |  |  |  |  | 0.25*<br>(0.14) |
| Share of 80+ |  |  |  |  |  |  | 0.17<br>(0.16) |
| Mean income |  |  |  |  |  |  | 0.28<br>(0.19) |
| Observations | 60,384 | 60,384 | 60,384 | 60,384 | 60,384 | 60,384 | 60,384 |
| R-squared | 0.24 | 0.24 | 0.24 | 0.24 | 0.24 | 0.24 | 0.25 |

**Notes:** See notes to Table 1. Column (1) reports the baseline specification (Column (5) in Table 1). Columns (2) and (3) change onset of treatment, Column (4) excludes the accommodation sector, Columns (5) and (6) perform placebo tests, and Column (7) reports the results from including additional controls.

**Figure A1:**
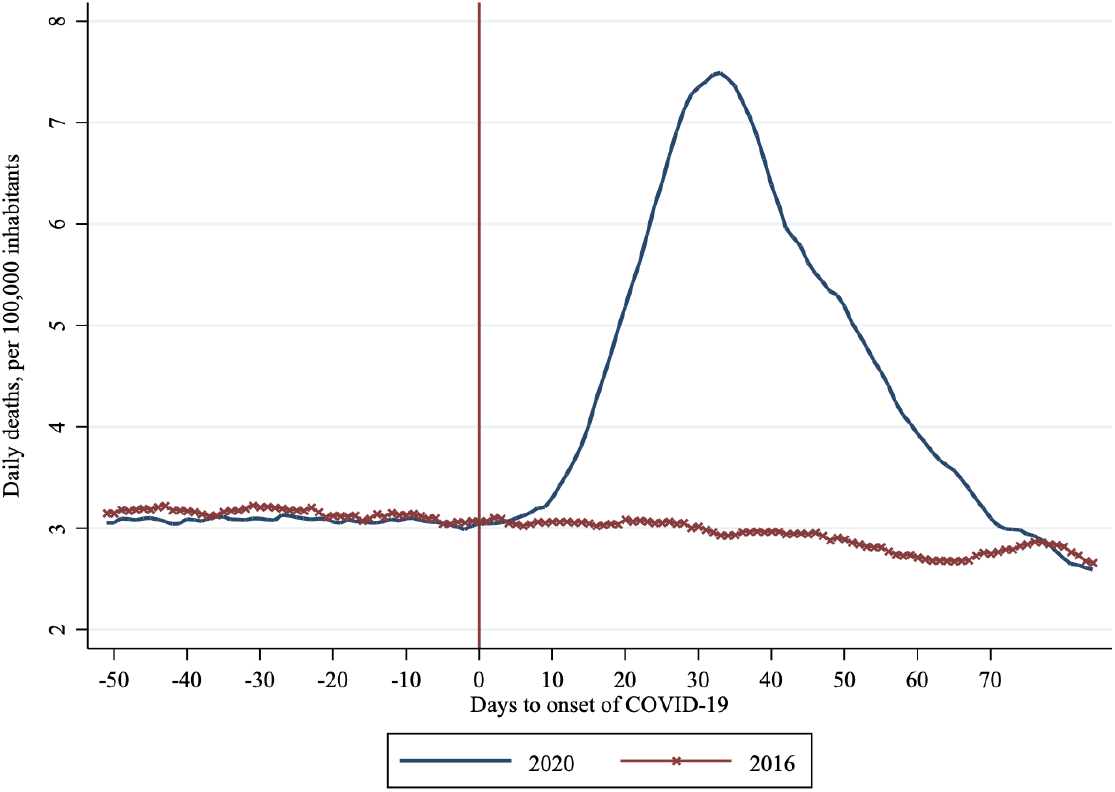
Daily mortality trends. **Notes:** This figure plots mortality trends in 2020 and for the synthetic control group, constructed following (Abadie et al., 2014).

**Figure A2:**
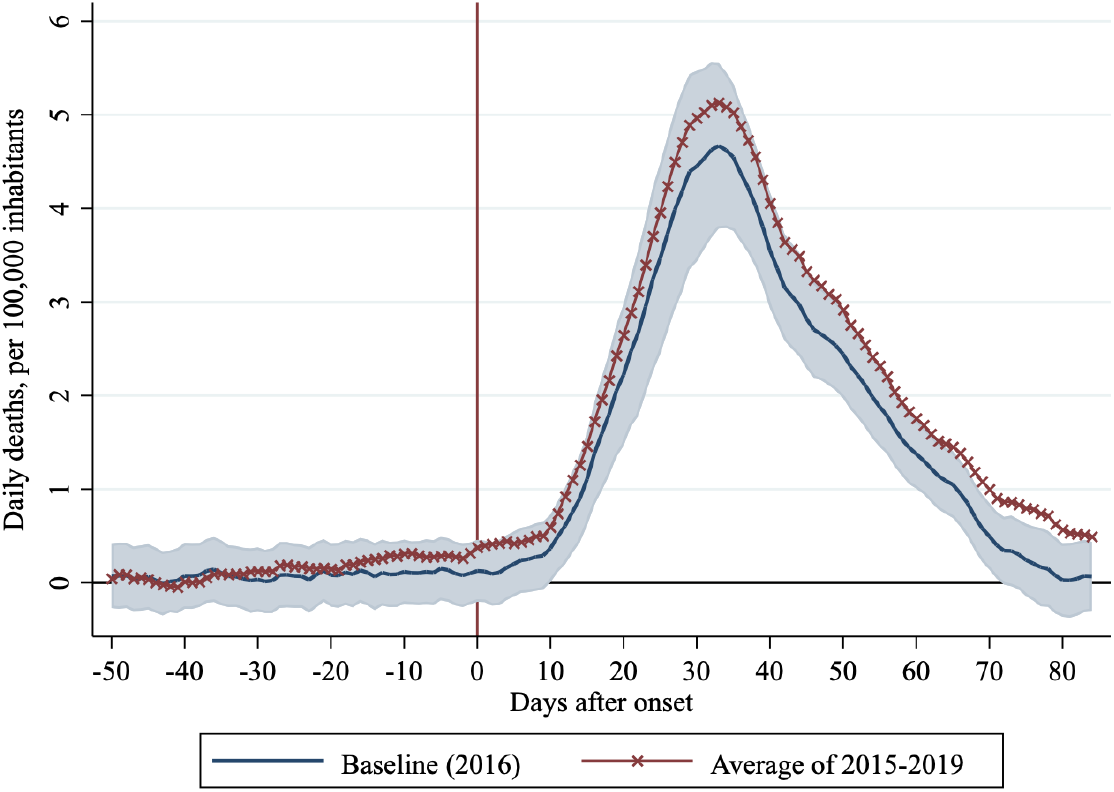
Robustness of the estimates to the control group. **Notes:** The figure shows the estimated effects of COVID-19 on mortality using 2016 as control group (in blue) and its 95% confidence bands. In red, it shows the estimated effects using the average mortality of years 2015-2019 as control group.

## Footnotes

1 Low-tech manufacturing activities include wearing apparel manufacturing, textiles manufacturing and beverages and tobacco manufacturing. High-tech manufacturing activities instead include chemicals and pharmaceuticals manufacturing as well as computers and electrical equipment manufacturing.

2 An exception is the work of Kaplan et al. (2020). These authors incorporate heterogeneity in contact by sector, which allow them to discuss the need for sector-specific policies.

3 Direct deaths refer to people dying of COVID-19, while indirect deaths refer to those dying for causes related to COVID-19, such as overcrowded hospitals (see The Economist, 2020). It should be noted, however, that COVID-19 may have also resulted in less deaths due to other causes, such as road accidents, accidents on the workplace and influenza. Hence, the number of indirect deaths is net of fewer deaths resulting from government social distancing policies.

4 We exclude agriculture and public administration from the analysis since data on the number of workers in these two sectors are not available at the municipality-level.

5 To construct the index we use data on the number of employees in each sector from ISTAT (2020*a*) as well as sector-specific scores in the propensity towards digital labor. Such scores are assigned by Manyika et al. (2015) and are reported in Table 3. The index is constructed computing the weighted sum of employment in sectors that stay open, using the scores of Manyika et al. (2015) as weights, and dividing it by the overall employment level (in open sectors). For the definition of open sectors, we rely on the list issued by the government (see Section 2.3 below for more details). Using the scores of Dingel and Neiman (2020) leads to a very similar series.

6 A few exceptions were granted. Restaurants and bars could offer takeaway and delivery, while non-essential production activities were allowed to operate if they were supplying goods to firms in essential sectors.

7 To pool municipalities into local labor markets we use the 2011 concordance table provided by ISTAT. Except if otherwise specified, all the variables that we collect are at the municipality-level. We compute their local labor market equivalent using the categorization by ISTAT.

8 Data on the number of workers disaggregated at the level of the local labor market is only available at the 3-digit level, while the government defined the sectors that had to close down using the 5-digit classification. To construct our business shutdowns variable, we source country-level data on the number of workers in each 5-digit sector and construct a measure of the share of suspended employment in each 3-digit sector at the national level, which we then use to compute the employment share in shutdown sectors in each local labor market.

9 Lauer et al. (2020) and Linton et al. (2020) study COVID-19 incubation and find a lag of about 5 days between infection and incubation. SARS-CoV-2 Surveillance Group (2020) follows a panel of COVID-19 deaths in Italy and finds a median time of 10 days between the onset of symptoms and death. We account for a minimum lag of 7 days between the implementation of the policy and its effects, but our results are robust to different choices (see Table A3 in the Appendix).

10 The choice of using mortality in 2016 as counterfactual follows from a visual inspection of the data (see Figure 1 above and Figure A1 in the Appendix) and is also confirmed when using the synthetic control group method of Abadie et al. (2010, 2014), which assigns unit weight to the year 2016. Our results are robust to other choices of counterfactual (see Figure A2).

11 We opt for population analytical weights due to the sensible differences in population size observed across local labor markets and because we will use the estimated coefficients to perform some back-of-the-envelope calculations on the overall mortality effects of COVID-19 (see Section 3.2 below). Our estimates are however robust to not weighting for population (results available upon request).

12 Education may affect health behavior (see Galama et al., 2018) and attitudes towards social-distancing practices (Adda, 2016). Moreover, the level of education typically correlates with underlining health conditions (Case and Deaton, 2017; Chetty et al., 2016), and Yang et al. (2020) have shown that patients with co-morbidities have a higher chance of dying from COVID-19. Unfortunately, data on health conditions at the municipality/local-labor-marketlevel are not available.

13 Several studies have shown long-term exposure to particulate matters such as PM10 and PM2.5 to increase health risks (for a review, see World Health Organization, 2003), while Wu et al. (2020); Conticini et al. (2020); Becchetti et al. (2020) have found a positive link between PM10 and COVID-19-induced mortality. Since data availability on air pollution is an issue, we use provincial-level data on the number of days in a year in which the level of PM10 is above the legal limit of 50 *mg/m*^3^ and match it to local labor markets. The coefficient that we estimate is positive but not significant. This may be due to the fact that the variable we use as proxy has limited variation.

14 Markowitz et al. (2019) have shown that higher employment rates are associated with a higher flu incidence, while Adda (2016); Harris (2020) have shed light of the important role of commuting for the spread of infectious diseases. We therefore control for both these factors. We also account for the share of residents enrolled at university to control for the contemporaneous government’s policy of moving higher education learning online. Finally, we control for the share of the elderly, since age is an important determinant of COVID-19 lethality (SARS-CoV-2 Surveillance Group, 2020), and for mean income because low-income agents tend to have worse health (Case and Deaton, 2017; Chetty et al., 2016), and Yang et al. (2020) have shown that patients with co-morbidities have a higher chance of dying from COVID-19.

15 In a companion paper (Ciminelli and Garcia-Mandicó, 2020), we extensively document the large spatial heterogeneity in the unfolding of the epidemic. Communities close to the epidemic epicentre (usually defined as the towns of Codogno and Alzano Lombardo in the Lombardy region) displayed much higher death rates relative to those far from it when the business shutdown policies were implemented.

16 In practice, the additional regressor that we include in the specification is as follows: 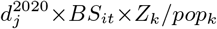, where the subscript *k* denotes province *k*; *Z*_*k*_ measures cumulative cases up to the day in which the policy is implemented; *pop*_*k*_ is population size; and the rest is as in Equation (3) above. We use data on cases at the provincial-level (rather than at the local-labor-market-level) since official data on detected cases from Protezione Civile (2020) are only available at this level of disaggregation. Hence, our measure of timeliness is provincerather than local-labor-marketvarying.

17 These calculations take into account both that business shutdowns are relatively more effective if implemented earlier in the epidemic curve and that, given their effectiveness in terms of daily lives saved, they lead to more saved lives if they are in place for longer.

18 Except for manufacturing, which we divide between high- and low-tech and trade, which we divide between retail and wholesale, we follow the 1-digit categorization of the ISIC Rev 4 classification. The decision to divide the manufacturing and trade industries in sub-sectors follow from the fact that these sub-sectors display different propensities towards digital labor (see more below).

19 Using Italy’s data, Navaretti et al. (2020) compute the GDP loss that would occur if each industry were to be closed for one year. Of the 12 industries that would cause the most significant costs, 11 belong to the 5 sectors that were most affected by the shutdowns.

## References

Abadie, A., Diamond, A. and Hainmueller, J. (2010), ‘Synthetic Control Methods for Comparative Case Studies: Estimating the Effect of California’s Tobacco Control Program’, Journal of the American Statistical Association 105(490), 493–505.

Abadie, A., Diamond, A. and Hainmueller, J. (2014), Synth: Stata Module to Implement Synthetic Control Methods for Comparative Case Studies, Working paper, Boston College Department of Economics.

Acemoglu, D. and Restrepo, P. (2020), ‘Robots and Jobs: Evidence from US Labor Markets’, Journal of Political Economy 128(6), 2188–2244.

Adda, J. (2016), ‘Economic Activity and the Spread of Viral Diseases: Evidence from High Frequency Data’, The Quarterly Journal of Economics 131(2), 891–941.

Allcott, H. and Keniston, D. (2018), ‘Dutch Disease or Agglomeration? the Local Economic Effects of Natural Resource Booms in Modern America’, The Review of Economic Studies 85(2), 695–731.

Atkeson, A. (2020), What Will Be the Economic Impact of COVID-19 in the US? Rough Estimates of Disease Scenarios, Working Paper 26867, National Bureau of Economic Research.

Becchetti, L., Conzo, G., Conzo, P. and Salustri, F. (2020), Understanding the Heterogeneity of Adverse COVID-19 Outcomes: the Role of Poor Quality of Air and Lockdown Decisions, Working Paper 3572548, SSRN.

Boniol, M., McIsaac, M., Xu, L., Wuliji, T., Diallo, K. and Campbell, J. (2019), Gender Equity in the Health Workforce: Analysis of 104 Countries, Technical report, World Health Organization.

Case, A. and Deaton, A. (2017), ‘Mortality and Morbidity in the 21st Century’, Brookings Papers on Economic Activity 2017(1), 397–476.

Chetty, R., Stepner, M., Abraham, S., Lin, S., Scuderi, B., Turner, N., Bergeron, A. and Cutler, D. (2016), ‘The Association Between Income and Life Expectancy in the United States, 2001-2014’, Journal of the American Medical Association 315(16), 1750–1766.

Chinazzi, M., Davis, J. T., Ajelli, M., Gioannini, C., Litvinova, M., Merler, S., y Piontti, A. P., Mu, K., Rossi, L., Sun, K. et al.. (2020), ‘The Effect of Travel Restrictions on the Spread of the 2019 Novel Coronavirus (COVID-19) Outbreak’, Science 368(6489), 395–400.

Ciminelli, G. and Garcia-Mandicó, S. (2020), COVID-19 in Italy: An Analysis of Death Registry Data, Mimeo.

Conticini, E., Frediani, B. and Caro, D. (2020), ‘Can Atmospheric Pollution be Considered a Co-Factor in Extremely High Level of SARS-CoV-2 Lethality in Northern Italy?’, Environmental Pollution p. 114465.

Dave, D., Friedson, A. I., Matsuzawa, K. and Sabia, J. J. (forthcoming), ‘When Do Shelter-in-Place Orders Fight COVID-19 Best? Policy Heterogeneity Across States and Adoption Time’, Economic Inquiry.

David, H., Dorn, D. and Hanson, G. H. (2013), ‘The China Syndrome: Local Labor Market Effects of Import Competition in the United States’, American Economic Review 103(6), 2121–68.

Dingel, J. I. and Neiman, B. (2020), ‘How Many Jobs Can Be Done at Home?’, Journal of Public Economics 189.

Eichenbaum, M. S., Rebelo, S. and Trabandt, M. (2020), The Macroeconomics of Epidemics, Working Paper 26882, National Bureau of Economic Research.

Fang, H., Wang, L. and Yang, Y. (2020), Human Mobility Restrictions and the Spread of the Novel Coronavirus (2019-ncov) in China, Working Paper 26906, National Bureau of Economic Research.

Ferguson, N., Laydon, D., Nedjati Gilani, G., Imai, N., Ainslie, K., Baguelin, M., Bhatia, S., Boonyasiri, A., Cucunuba Perez, Z., Cuomo-Dannenburg, G. et al.. (2020), Report 9: Impact of Non-Pharmaceutical Interventions (NPIs) to Reduce COVID-19 Mortality and Healthcare Demand, Technical report, Imperial College London.

Friedson, A., McNichols, D., Sabia, J. and Dave, D. (2020), Did California’s Shelter-in-Place Order Work? Early Coronavirus-Related Public Health Effects, Working Paper 26992, National Bureau of Economic Research.

Galama, T. J., Lleras-Muney, A. and van Kippersluis, H. (2018), The Effect of Education on Health and Mortality: A Review of Experimental and Quasi-Experimental Evidence, Working Paper 24225, National Bureau of Economic Research.

Harris, J. E. (2020), The Subways Seeded the Massive Coronavirus Epidemic in New York City, Working Paper 27021, National Bureau of Economic Research.

ISTAT (2011), ‘Sistemi Locali del Lavoro, 2011’. Data retrieved on April 12, 2020, www.istat.it/sistemi-locali-del-lavoro.

ISTAT (2018), ‘Condizioni Socio-Economiche delle Famiglie’. Data retrieved on April 12, 2020, https://www.istat.it/it/archivio/190365.

ISTAT (2019), ‘Dati Ambientali nelle Città. Ambiente Urbano’. Data retrieved on April 12, 2020, https://www.istat.it/it/archivio/55771.

ISTAT (2020a), ‘Atlante Statistico dei Comuni’. Data retrieved on April 15, 2020, http://asc.istat.it/asc_BL/.

ISTAT (2020b), ‘Decessi del 2020. Dataset Analitico con i Decessi Giornalieri.’. Data retrieved on June 4, 2020, https://www.istat.it/it/archivio/242055.

ISTAT (2020c), ‘Indicatori Demografici. Popolazione Residente al 1 Gennaio.’. Data retrieved on April 12, 2020, https://www.istat.it/it/popolazione-e-famiglie?dati.

ISTAT (2020d), ‘A Misura di Comune’. Data retrieved on April 15, 2020, https://www.istat.it/it/archivio/220004.

Jones, C. J., Philippon, T. and Venkateswaran, V. (2020), Optimal Mitigation Policies in a Pandemic: Social Distancing and Working from Home, Working Paper 26984, National Bureau of Economic Research.

Juranek, S. and Zoutman, F. (2020), The Effect of Social Distancing Measures on the Demand for Intensive Care: Evidence on COVID-19 in Scandinavia, Working Paper 8262, CESifo.

Kaplan, G., Moll, B. and Violante, G. (2020), Pandemics According to HANK, Powerpoint presentation, LSE.

Khain, E. (2020), Two-Level Modeling of Quarantine, Preprint 2005.01505, arXiv.

Lauer, S. A., Grantz, K. H., Bi, Q., Jones, F. K., Zheng, Q., Meredith, H. R., Azman, A. S., Reich, N. G. and Lessler, J. (2020), ‘The Incubation Period of Coronavirus Disease 2019 (COVID-19) from Publicly Reported Confirmed Cases: Estimation and Application’, Annals of Internal Medicine 172(9).

Lewandowski, P. et al.. (2020), Occupational Exposure to Contagion and the Spread of COVID-19 in Europe, Discussion Paper 13277, Institute of Labor Economics (IZA).

Linton, N. M., Kobayashi, T., Yang, Y., Hayashi, K., Akhmetzhanov, A. R., Jung, S.-m., Yuan, B., Kinoshita, R. and Nishiura, H. (2020), ‘Incubation Period and Other Epidemiological Characteristics of 2019 Novel Coronavirus Infections with Right Truncation: a Statistical Analysis of Publicly Available Case Data’, Journal of Clinical Medicine 9(2), 538.

Manyika, J., Ramaswamy, S., Khanna, S., Sarrazin, H., Pinkus, G., Sethupathy, G. and Yaffe, A. (2015), Digital America: A Tale of the Haves and Have-Mores, Technical report, McKinsey Global Institute.

Markowitz, S., Nesson, E. and Robinson, J. J. (2019), ‘The Effects of Employment on Influenza Rates’, Economics & Human Biology 34, 286–295.

Moretti, E. (2010), Local Labor Markets, Working Paper 15947, National Bureau of Economic Research.

Muellbauer, J. and Aron, J. (2020), Measuring Excess Mortality: The Case of England during the COVID-19 Pandemic, Working Paper 2020-11, Institute for New Economic Thinking.

Navaretti, G. B., Calzolari, G., Dossena, A., Lanza, A. and Pozzolo, A. F. (2020), ‘In and Out Lockdowns: Identifying the Centrality of Economic Activities’, Covid Economics 17, 189–204.

Pedersen, M. G. and Meneghini, M. (2020), Quantifying Undetected COVID-19 Cases and Effects of Containment Measures in Italy, Preprint, ResearchGate.

Protezione Civile (2020), Covid-19 italia - monitoraggio della situazione, Technical report. URL: http://opendatadpc.maps.arcgis.com

Rajan, R. G. and Zingales, L. (1998), ‘Financial Dependence and Growth’, The American Economic Review 88(3), 559–586.

SARS-CoV-2 Surveillance Group (2020), Characteristics of SARS-CoV-2 Patients Dying in Italy. Report Based on Available Data on April 2nd, 2020, Technical report, Istituto Superiore di Sanità.

The Economist (2020), Deaths from Cardiac Arrest Have Surged in New York City., Online article.

World Health Organization (2003), Health Aspects of Air Pollution with Particulate Matter, Ozone and Nitrogen Dioxide: Report on a WHO Working Group, Bonn, Germany 13-15 January 2003, Technical report, WHO Regional Office for Europe.

Wu, X., Nethery, R. C., Sabath, B. M., Braun, D. and Dominici, F. (2020), Exposure to air pollution and covid-19 mortality in the united states, Working paper.

Yang, J., Zheng, Y., Gou, X., Pu, K., Chen, Z., Guo, Q., Ji, R., Wang, H., Wang, Y. and Zhou, Y. (2020), ‘Prevalence of Comorbidities in the Novel Wuhan Coronavirus (COVID-19) Infection: A Systematic Review and Meta-Analysis’, International Journal of Infectious Diseases.

